# Adiposity-Associated Monocyte Costimulatory Programming in Rheumatoid Arthritis Identified by Single-Cell Transcriptomics

**DOI:** 10.64898/2026.06.09.26355275

**Authors:** Shalini N. Swamy, Hua Zhong, Kylie Williams, Joan T. Merrill, Kurt Zimmerman, Beatriz Y. Hanaoka

**Affiliations:** Section of Rheumatology, Allergy and Immunology, Department of Medicine, College of Medicine, University of Oklahoma Health Sciences, Oklahoma City, OK, USA; Department of Biostatistics and Epidemiology, Hudson College of Public Health, University of Oklahoma Health Sciences, Oklahoma City, OK, USA; Molecular Analysis and Cellular Imaging (MACI) Core, Center for Geroscience, University of Oklahoma Health Campus, Oklahoma City, OK, USA; Arthritis and Clinical Immunology Research Program, Oklahoma Medical Research Foundation, Oklahoma City, OK, USA; Department of Medicine, College of Medicine, University of Oklahoma Health Sciences, Oklahoma City, OK, USA

**Author notes:** **Corresponding Author:** Beatriz Y. Hanaoka, MD, MSc, Division of Rheumatology, University of Oklahoma Health Sciences Center, Oklahoma City, OK, USA.

## Abstract

**Background:** Rheumatoid arthritis (RA) is a chronic systemic inflammatory disease which can lead to progressive disability and damage to multiple organs. Obesity is associated with higher disease activity in RA and inadequate long-term outcomes, so better understanding of mechanisms linking adiposity to immune dysregulation might help to refine optimal treatments. Monocytes are important contributors to immune activation in RA through antigen presentation and costimulatory signaling. We hypothesized that adiposity enhances monocyte costimulatory programming in RA, thereby promoting adaptive immune activation.

**Methods:** Single-cell RNA sequencing was performed using the 10x Genomics Flex platform on purified circulating monocytes from 31 donors (16 RA participants fulfilling 2010 ACR/EULAR classification criteria and 15 non-RA controls) generating transcriptomic profiles for approximately 135,599 monocytes. Donor-level pathway enrichment scores were calculated for predefined immune activation pathways including antigen processing and presentation, interferon signaling, and regulation of T-cell costimulation. Analyses were performed at the donor level to avoid cell-level pseudoreplication. Associations with disease status and body mass index were evaluated using factorial linear models and Spearman correlation analyses.

**Results:** Single-cell transcriptomic profiling identified classical, intermediate-like, non-classical, and interferon-responsive monocyte populations. RA was associated with enrichment of antigen processing and presentation programs in circulating monocytes (p=0.0106), indicating a primed antigen-presenting state. In contrast, regulation of T-cell costimulation pathway enrichment did not differ by RA status alone. However, within RA participants, higher BMI was associated with increased enrichment of monocyte T-cell costimulatory pathways (Spearman ρ=0.56, p=0.0248), unlike in non-RA controls. Gene-level analyses demonstrated strong baseline expression of CD86, while ICOSLG and TNFSF4 transcripts were expressed at low levels overall, consistent with inducible costimulatory signaling programs.

**Conclusions:** These findings support a model in which metabolic dysregulation amplifies monocyte-mediated immune activation and may contribute to worsened disease outcomes in RA.

## Introduction

Rheumatoid arthritis (RA) is a chronic systemic autoimmune disease characterized by persistent synovial inflammation, progressive joint damage, and potential for substantial functional disability. Obesity is recognized as an important modifier of immunologic processes relevant to RA disease activity with multiple studies implicating associations between increased adiposity and higher disease activity and reduced likelihood of remission. (1-4). Mechanisms linking obesity-associated inflammation to immune dysregulation in RA may be multifactorial, including both mechanical causes and direct induction of modified of immunologic processes. To the extent that the immunologic impacts of obesity on RA can be better defined, strategic modifications in treatment might improve outcomes.

Monocytes contribute to inflammatory amplification in RA through cytokine production, antigen presentation, and interactions with adaptive immune cells (5, 6). In addition to their established role in synovial inflammation, circulating monocytes integrate inflammatory cues derived from peripheral tissues and may contribute to obesity-associated immune activation Recent single-cell studies have illuminated substantial transcriptional heterogeneity within monocyte populations and suggest that inflammatory and interferon-responsive activation programs extend across traditional subset boundaries, supporting a transcriptionally defined activation-state framework rather than rigid phenotypic classifications (5-7).

Obesity is characterized by chronic low-grade inflammation and altered myeloid-cell homeostasis (8, 9). Monocytes are responsive to inflammatory environmental cues and may amplify immune activation through antigen presentation and costimulatory signaling. Some observations suggest that both RA and obesity may contribute to monocyte activation. In RA, expansion of circulating CD14^+^CD16^+^ intermediate monocytes have been associated with disease activity (10), and obesity has been linked to enhanced myelopoiesis and monocytosis (11). These observations suggest that RA and obesity may converge on monocyte-lineage activation. However, whether obesity-associated monocyte changes in RA reflect shifts in canonical monocyte subsets or broader transcriptional activation states remain unclear.

We hypothesized that adiposity is associated with altered monocyte activation states in RA, and used single-cell RNA sequencing (scRNA-seq) to define monocyte activation programs in people with and without RA over a range of body mass index (BMI). Using donor-level pathway enrichment analyses, we evaluated associations between adiposity and monocyte inflammatory and immune activation programs, focusing on antigen presentation, inflammatory, interferon-responsive, and costimulatory pathways.

## Methods

### Study Participants

Peripheral blood samples and clinical data were obtained from participants with RA and non-RA controls enrolled through Institutional Review Board-approved observational studies. All participants provided written informed consent prior to enrollment. All participants provided written informed consent prior to enrollment and sample collection. All participants provided written informed consent prior to enrollment. Eligible participants were adults aged 18 years or older. RA participants fulfilled the 2010 American College of Rheumatology/European Alliance of Associations for Rheumatology classification criteria for RA (12), had disease duration ≥6 months, and were receiving stable therapy for at least 8 weeks prior to enrollment. Non-RA controls had no known diagnosis of RA or other systemic autoimmune inflammatory disease.

Body mass index (BMI) was calculated as weight in kilograms divided by height in meters squared (kg/m^2^). For descriptive analyses, participants were categorized as lean (BMI <25.0 kg/m^2^), overweight (BMI 25.0–29.9 kg/m^2^), or obese (BMI ≥30.0 kg/m^2^). Clinical characteristics collected included disease duration, rheumatoid factor (RF) and anti-cyclic citrullinated peptide (anti-CCP) antibody status, and medication use at study enrollment. Disease activity was assessed using the 3-component Disease Activity Score in 28 joints with C-reactive protein (DAS28-CRP[3]) (13), calculated from tender joint count, swollen joint count, and CRP. Medications were categorized as glucocorticoids, conventional synthetic disease-modifying antirheumatic drugs (csDMARDs), and biologic or targeted synthetic disease-modifying antirheumatic drugs (bDMARDs/tsDMARDs).

### Monocyte Isolation and Single-Cell RNA Sequencing

Single-cell RNA sequencing (scRNA-seq) libraries were generated using the 10x Genomics Chromium Flex Gene Expression platform according to the manufacturer’s protocol. Cell viability was assessed by trypan blue exclusion prior to processing, then assessed following fixation as well using propidium iodide staining. Single-cell suspensions were prepared for probe hybridization, barcode generation, and library construction using the Chromium X Controller workflow. Libraries underwent quality-control assessment and were sequenced on an Illumina platform using paired-end sequencing.

scRNA-seq was initially performed on purified circulating monocytes from 32 donors. One donor sample failed quality-control filtering and was excluded from downstream analyses, resulting in a final analytic cohort of 31 donors (16 RA participants and 15 non-RA controls). Following quality-control filtering, 135,599 cells were retained for downstream analyses.

### Single-Cell Data Processing and Clustering

Sequencing data underwent quality-control filtering, normalization, dimensionality reduction, and clustering using Seurat in R. Cells with low transcript counts or high mitochondrial transcript percentages were excluded. Uniform manifold approximation and projection (UMAP) was used for visualization of transcriptional populations. Monocyte populations were annotated using established marker genes, including CD14, FCGR3A, S100A8, S100A12, IFI44L, ISG15, and MX1.

### Pathway Enrichment Analyses

To avoid pseudoreplication, pathway enrichment analyses were summarized at the donor level rather than analyzed at the individual cell level. Predefined immune activation pathways relevant to the study hypothesis were evaluated, including antigen processing and presentation, interferon signaling, inflammatory response, and regulation of T-cell costimulation. Disease- and adiposity-stratified analyses were performed using donor-level enrichment scores.

### Statistical Analysis

Statistical analyses were performed in R (v4.0+). Associations between body mass index (BMI) and pathway enrichment scores were evaluated using Spearman correlation analyses. Disease- and adiposity-associated effects were additionally evaluated using factorial linear models. Two-sided p values <0.05 were considered statistically significant.

## Results

### Participant characteristics

Participant characteristics are summarized in Table 1. The final analytic cohort consisted of 31 participants, including 16 individuals with rheumatoid arthritis (RA) and 15 non-RA controls. The median age was 52.5 years (IQR 42.5–60.0) in the RA group and 36.0 years (IQR 31.0–52.0) among controls. Most participants were female (93.8% of RA participants and 80.0% of controls). BMI distributions were similar between groups, with approximately one-third of participants classified as obese.

RA participants had a median disease duration of 7.5 years (IQR 4.5–13.0). Among the 14 RA participants with available serologic data, 7 (50.0%) were rheumatoid factor (RF) positive and 5 (35.7%) were anti-cyclic citrullinated peptide (anti-CCP) antibody positive. Disease activity was generally low, with a median DAS28-CRP(3) of 1.75 (IQR 1.35–3.66), indicating remission at the cohort level overall. Regarding treatment exposure, 4 (25.0%) participants were receiving glucocorticoids, 10 (62.5%) conventional synthetic disease-modifying antirheumatic drugs (csDMARDs), and 8 (50.0%) biologic or targeted synthetic DMARDs (b/tsDMARDs).

| Characteristic | RA (n=16) | Non-RA Controls (n=15) |
| --- | --- | --- |
| Age, years (IQR) | 52.5 (42.5-60.0) | 36.0 (31.0-52.0) |
| Female sex, n (%) | 15 (93.8%) | 12 (80%) |
| BMI, kg/m<sup>2</sup> (IQR) | 26.8 (24.2–40.5) | 27.1 (23.9–36.4) |
| Lean, n (%) | 5 (31.3) | 6 (40.0) |
| Overweight, n (%) | 6 (37.5) | 3 (20.0) |
| Obese, n (%) | 5 (31.3) | 6 (40.0) |
| RA disease duration, years | 7.5 (4.5–13.0) | NA |
| RF positive, n (%)* | 7 (50.0) | NA |
| Anti-CCP positive, n (%)* | 5 (35.7) | NA |
| DAS28-CRP[3], (IQR)* | 1.745 (1.35-3.66) | NA |
| Glucocorticoid use, n (%) | 4 (25.0) | NA |
| cs DMARD use, n (%) | 10 (62.5) | NA |
| Biologic/tsDMARD use, n (%) | 8 (50.0) | NA |

\*Results were available for 14 of 16 RA participants.

### Single-cell RNA sequencing identifies transcriptionally distinct circulating monocyte populations

Following quality-control filtering, 135,599 monocytes were retained for downstream single-cell transcriptomic analyses. Analysis of circulating monocytes from 31 donors revealed transcriptionally distinct populations corresponding to classical, intermediate-like, and non-classical monocyte states, together with a discrete interferon (IFN)-responsive population (Figure 1A). These populations were defined by characteristic gene expression programs consistent with established monocyte subsets.

**Figure 1.**
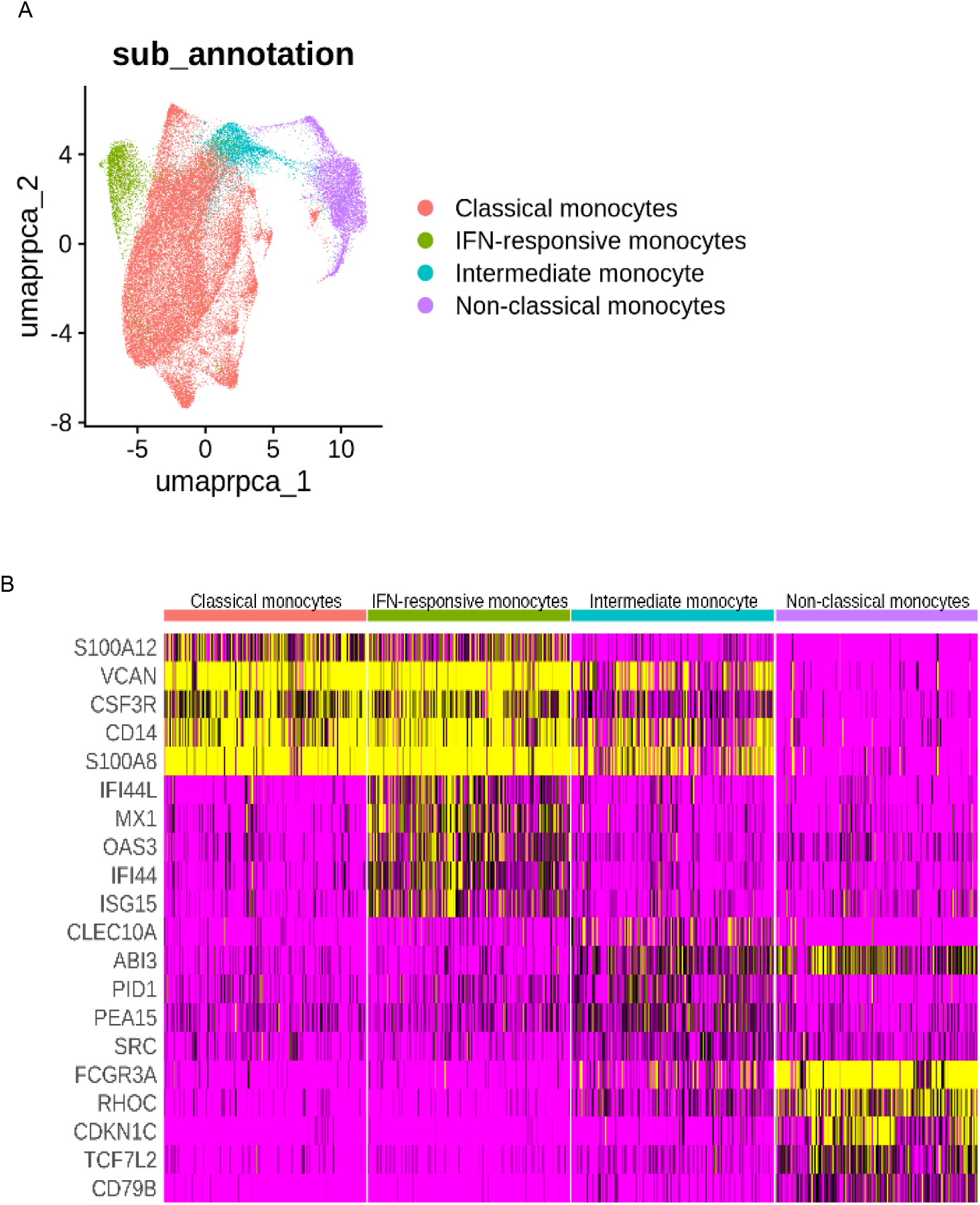
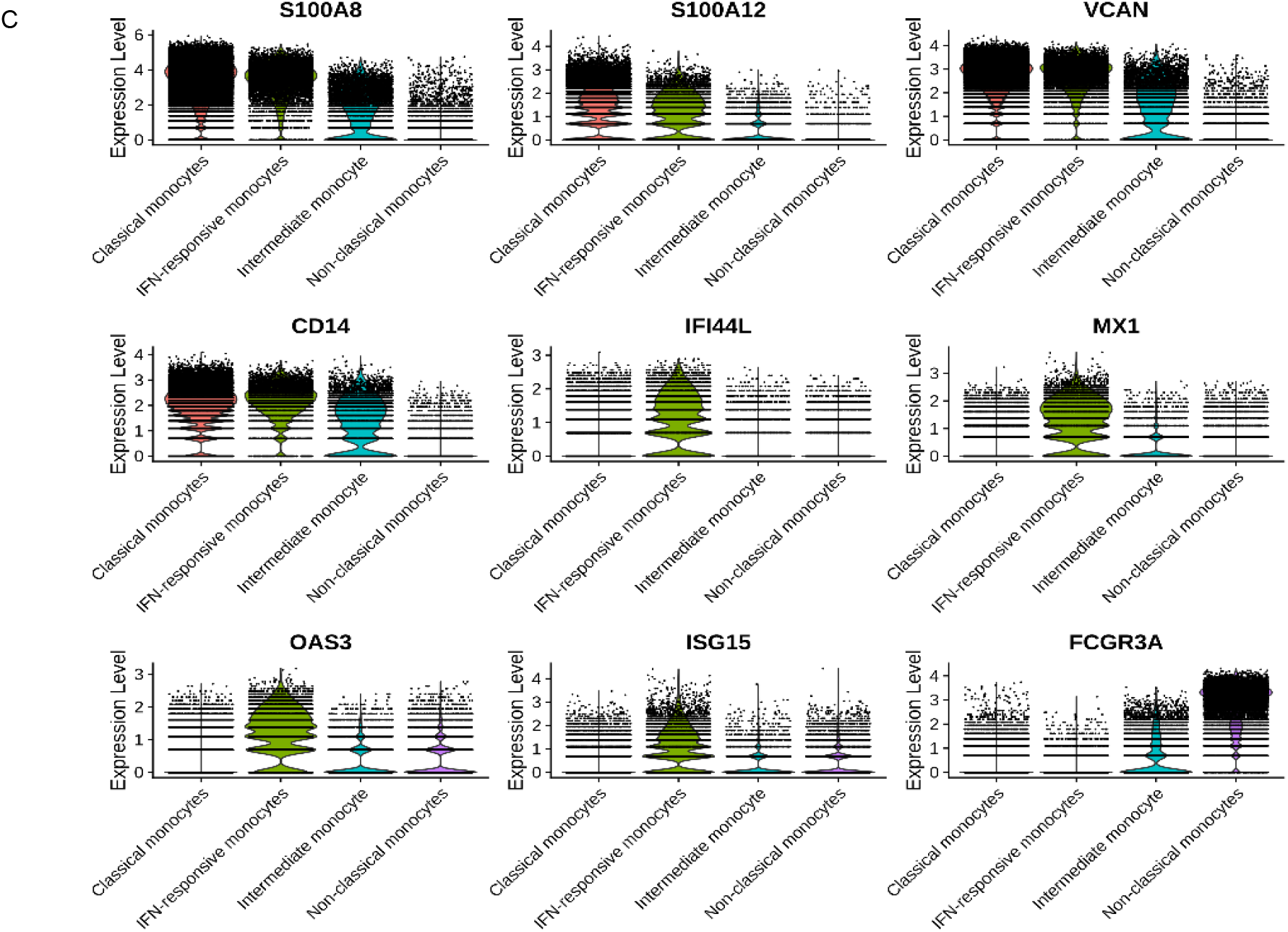
Single-cell transcriptomic profiling identifies transcriptionally distinct circulating monocyte populations. (A) Uniform manifold approximation and projection (UMAP) visualization of purified circulating monocytes from rheumatoid arthritis (RA) and non-RA participants across body mass index (BMI) categories demonstrating transcriptionally distinct monocyte populations. Clusters corresponded to classical, intermediate-like, non-classical, and interferon (IFN)-responsive monocyte states. (B) Heatmap of representative marker genes used for cluster annotation. Classical monocytes demonstrated enrichment of CD14, S100A8, S100A12, and VCAN, while non-classical monocytes were enriched for FCGR3A expression. The IFN-responsive cluster demonstrated elevated expression of interferon-stimulated genes including IFI44L, MX1, OAS3, and ISG15. (C) Representative gene-expression distributions across monocyte populations demonstrating transcriptional heterogeneity and IFN-responsive activation programs extending across canonical monocyte subset boundaries.

Cluster annotation was supported by canonical marker-gene expression patterns, including enrichment of CD14, S100A8, S100A12, and VCAN in classical monocytes and FCGR3A expression in non-classical monocytes (Figure 1B). A transcriptionally distinct IFN-responsive cluster demonstrated elevated expression of interferon-stimulated genes including IFI44L, MX1, OAS3, and ISG15 (Figure 1B-C). IFN-responsive transcriptional programs extended across canonical monocyte subset boundaries, supporting a transcriptionally defined activation-state framework rather than rigid phenotypic classification.

### RA is associated with enhanced monocyte antigen presentation pathway enrichment

To investigate immune activation programs relevant to RA pathogenesis, we performed donor-level pathway enrichment analyses using predefined immune activation pathways. Donor-level analyses were used to avoid pseudoreplication and permit disease- and adiposity-stratified comparisons.

Compared with non-RA participants, RA participants demonstrated significantly increased enrichment of antigen processing and presentation pathways (p=0.0106), consistent with enhanced antigen-presenting cell activation states in RA (Figure 2A). These findings support the presence of circulating monocyte activation programs characterized by increased antigen presentation capacity in RA.

**Figure 2.**
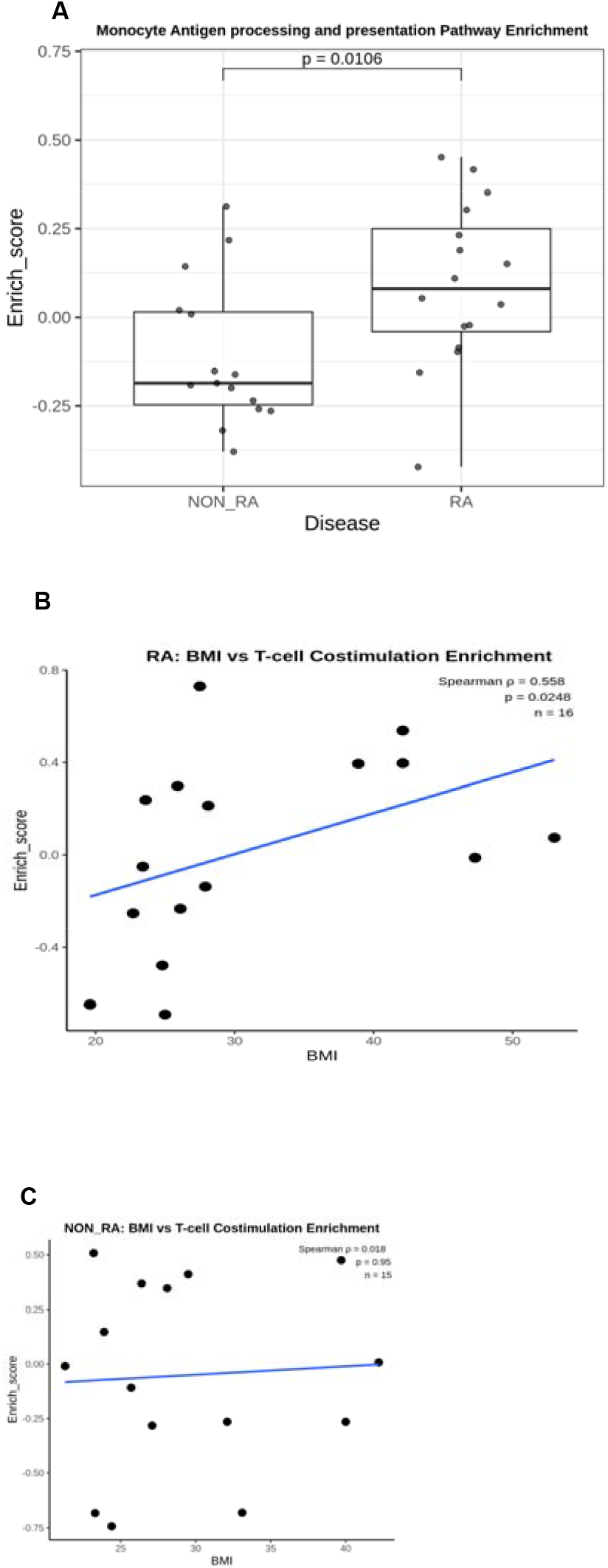
RA-associated antigen presentation and adiposity-associated monocyte costimulatory pathway enrichment. (A) Donor-level pathway enrichment analysis demonstrating increased antigen processing and presentation pathway enrichment in rheumatoid arthritis (RA) participants compared with non-RA controls. (B) Within RA participants, body mass index (BMI) was positively associated with monocyte regulation of T-cell costimulation pathway enrichment. (C) No significant association between BMI and monocyte regulation of T-cell costimulation pathway enrichment was observed among non-RA controls. Pathway enrichment analyses were summarized at the donor level to avoid pseudoreplication.

### Adiposity is associated with monocyte costimulatory pathway enrichment specifically within RA participants

Because obesity is associated with worsened RA disease activity and therapeutic responsiveness, we next evaluated relationships between adiposity and monocyte costimulatory activation programs. Regulation of T-cell costimulation pathway enrichment did not significantly differ according to RA status alone. However, within RA participants, body mass index (BMI) demonstrated a significant positive association with monocyte costimulatory pathway enrichment (Spearman ρ=0.56, p=0.0248), whereas no association was observed among non-RA controls (Figure 2B-C).

At the individual gene level, CD86 transcripts were broadly expressed across monocyte populations and demonstrated modest enrichment in RA, whereas ICOSLG and TNFSF4 expression was low at baseline, consistent with inducible regulation of costimulatory pathways in circulating monocytes. Together, these findings support an RA-context–specific association between adiposity and monocyte costimulatory programming.

## Discussion

In this study, single-cell transcriptomic profiling identified obesity-associated monocyte activation programs in rheumatoid arthritis (RA). We observed enhanced antigen processing and presentation pathway enrichment in RA monocytes, consistent with a primed antigen-presenting state, and identified a positive association between adiposity and monocyte costimulatory pathway enrichment within RA participants. These findings support a model in which obesity-associated metabolic dysregulation amplifies monocyte immunostimulatory programming within the inflammatory RA environment.

Monocytes are recognized as important mediators linking systemic metabolic dysfunction to adaptive immune activation. In RA, monocytes contribute to inflammatory amplification through cytokine production, antigen presentation, and costimulatory signaling. Our findings may be relevant to this framework by demonstrating that adiposity is associated with enhanced monocyte costimulatory programming within RA patients. In this small study, the phenomenon was not detected as a generalized feature of obesity alone, however this does not rule out the possibility that similar findings would emerge from a less immunologically activated population with a greater sample size. Either way, our data suggests that obesity-associated metabolic signals may interact with the inflammatory RA milieu to augment immune activation pathways relevant to disease persistence.

We identified a transcriptionally distinct interferon-responsive monocyte population characterized by expression of interferon-stimulated genes, including IFI44L, MX1, OAS3, and ISG15. IFN-responsive transcriptional programs extended across canonical monocyte subset boundaries, supporting a transcriptionally defined activation-state framework that complements traditional phenotypic classification. These findings are consistent with single-cell studies demonstrating substantial transcriptional and functional heterogeneity within circulating monocyte populations in inflammatory disease states (14).

We previously demonstrated that circulating monounsaturated fatty acids (MUFAs), including oleic acid, are associated with RA disease activity and abdominal adiposity independent of dietary intake (15). Together with the current transcriptomic findings, these observations raise the possibility that adiposity-associated metabolic alterations may contribute to monocyte immune conditioning in RA.

This pilot study has several limitations. The cohort size was small, and the analyses were exploratory and hypothesis-generating. Functional validation of the identified costimulatory pathways was not performed within this study and will require future mechanistic investigation. In addition, pathway enrichment analyses were performed using circulating monocytes and may not fully reflect tissue-level immune activation within the synovium or adipose tissue microenvironment. The control group was younger than the RA cohort, and age-related differences in immune cell transcriptional programs may have contributed to some observed differences between groups. Finally, RA is a clinically and immunologically heterogeneous disease, and participants were receiving a variety of therapies that may have influenced monocyte transcriptional programs.

## Data Availability

All data produced in the present study are available upon reasonable request to the authors.

## Declarations

### Ethics approval and consent to participate

The study was conducted in accordance with the Declaration of Helsinki and approved by the University of Oklahoma Health Sciences Center Institutional Review Board (IRB #16066).

Additional samples were obtained from participants enrolled in the Oklahoma Cohort of Rheumatic Diseases (IRB #09-21). All participants provided written informed consent.

### Consent for publication

Not applicable.

### Availability of data and materials

The datasets generated and/or analyzed during the current study are not publicly available because of participant privacy considerations but are available from the corresponding author upon reasonable request.

## Competing interests

The authors declare that they have no competing interests.

## Funding

This work was supported by the Rheumatology Research Foundation Bridge Award and the Presbyterian Health Foundation Bridge Award (PI: Beatriz Y. Hanaoka).

Additional support for research infrastructure and core resources was provided by the Oklahoma Nathan Shock Center of Excellence in the Biology of Aging and the Oklahoma Center for Geroscience and Healthy Brain Aging through the National Institute of General Medical Sciences Centers of Biomedical Research Excellence (COBRE) grant P20GM134973.

Research reported in this publication also utilized resources supported by an Institutional Development Award (IDeA) from the National Institute of General Medical Sciences under award number U54GM104938, by the National Institute of Arthritis and Musculoskeletal and Skin Diseases under award number P30AR073750, and by the National Institute of Allergy and Infectious Diseases under award number UM1AI144292 of the National Institutes of Health.

The funding sources had no role in study design, data collection, data analysis, interpretation of data, manuscript preparation, or the decision to submit the manuscript for publication. The content is solely the responsibility of the authors and does not necessarily represent the official views of the National Institutes of Health.

## Authors’ Contributions

BH conceived and designed the study; supervised participant recruitment and clinical phenotyping; interpreted the data; and drafted the manuscript. SNS assisted with study design, processed and prepared samples for single-cell RNA sequencing, contributed to data interpretation, and critically revised the manuscript. HZ performed bioinformatic analyses and contributed to data interpretation. KW performed library preparation and assisted with single-cell RNA sequencing data generation and quality control. JTM contributed participant samples and clinical data and critically reviewed the manuscript. KZ supervised the bioinformatic analyses, contributed to data interpretation, and critically revised the manuscript. All authors reviewed and approved the final manuscript.

## Acknowledgements

The authors thank all study participants for their contribution to this research. We also acknowledge the Oklahoma Medical Research Foundation (OMRF) Genomics Core for sequencing support and the Oklahoma Clinical and Translational Science Institute (OCTSI) study coordination and regulatory teams for their assistance with participant recruitment, study coordination, and regulatory oversight. We thank Wade DeJager for assistance with clinical data collection and study coordination at OMRF.

## Manuscript preparation / AI disclosure

Portions of the manuscript text were drafted and edited with the assistance of a Large Language Model (ChatGPT, OpenAI, San Francisco, CA, USA). All study design, data analysis, interpretation, and final text were reviewed and verified by the authors, who take full responsibility for the content.

## Notes

### Competing Interest Statement

The authors have declared no competing interest.

### Author Declarations

The study was approved by the University of Oklahoma Health Sciences Center Institutional Review Board (IRB #16066). Additional samples were obtained from participants enrolled in the Oklahoma Cohort of Rheumatic Diseases (IRB #09-21). All participants provided written informed consent.

